# Representation of Older Adults and Women in Randomized Trials of Non-Invasive Imaging for Chest Pain

**DOI:** 10.1101/2025.04.23.25326261

**Authors:** Phillip Lim, Tansu Eris, Leslee J. Shaw, Laura Gelfman, Annetine Gelijns, Alan Moskowitz, Emilia Bagiella, Fay A. Lin, Deepak L. Bhatt, Gregg Stone, R. Sean Morrison, David Cohen, Michael Nanna, Karen Alexander, Krishna K. Patel

## Abstract

**Background:** Non-invasive imaging is widely used both for initial diagnosis and to guide management of ischemic heart disease (IHD). Older adults and women with IHD may have different responses to imaging as well as to treatments and outcomes that follow compared with younger adults and men. We aimed to study the representation of older adults and women in randomized controlled trials (RCT) of non-invasive imaging among patients with acute and stable chest pain.

**Methods:** We conducted a systematic search to identify RCTs evaluating non-invasive, imaging-guided diagnosis and management for IHD that were published before September 1, 2023. Participation-to-Prevalence Ratio (PPR) was estimated for women and age subgroups of <65, 65-74, ≥75 years. PPR of <0.8, 0.8-1.2, and >1.2 indicated underrepresentation, appropriate representation, and overrepresentation, respectively.

**Results:** Among 53 RCTs, sex and age breakdown were available in 53 (n=55,893) and 21 trials (n=35,503), respectively. The median age across all trials was 57.4 years [IQR: 55.0– 60.2]. Participants aged <65 years were overrepresented with a median PPR 2.13 [IQR: 1.73– 2.43], while those aged 65–74 years and ≥75 years were underrepresented with median PPRs of 0.74 [IQR: 0.56–0.83] and 0.21 [IQR: 0.11–0.33], respectively. Women were adequately represented with a median PPR of 1.2 [1.06–1.32].

**Conclusion:** While women were appropriately represented, adults 65 years or older, especially those ≥75 years, were under-represented in these trials. Future RCTs on non-invasive imaging should target enrollment of older adults to ensure generalizability of results to this growing population.

**CLINICAL PERSPECTIVE:** In a systematic review of 53 randomized controlled trials of non-invasive imaging for chest pain published before September 1, 2023 (n=55,893 participants), adults aged 65 years and older, especially those aged 75 years and above, were significantly underrepresented, whereas women had representation proportional to prevalence estimates. These findings highlight an urgent need to increase enrollment of older adults in future imaging trials to ensure broader applicability and relevance of study results.

GRAPHICAL ABSTRACT:
Representation of Older Adults and Women in Randomized Controlled Trials of Non-Invasive Imaging for Ischemic Heart Disease

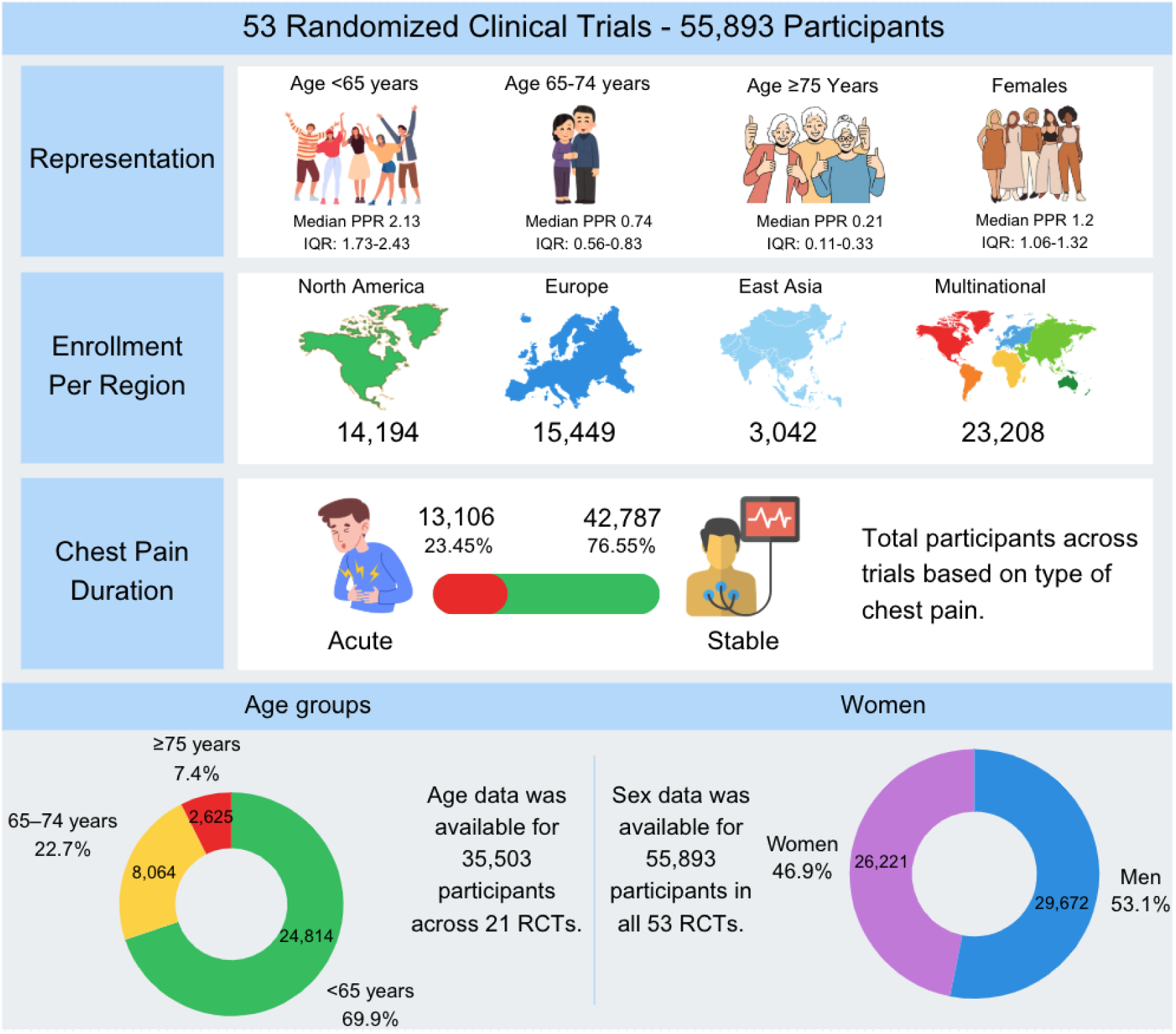

## Introduction

Ischemic heart disease (IHD) remains the leading cause of cardiovascular morbidity and mortality worldwide ^1,2^, accounting for substantial global health and economic burden. Non-invasive imaging modalities such as nuclear stress imaging, stress echocardiography, stress cardiac magnetic resonance imaging and coronary computed tomography angiography (CCTA) play a central role in the diagnosis and management of IHD, particularly among patients presenting with symptoms of ischemia such as chest pain, dyspnea and exertional intolerance.

As global life expectancy continues to rise, the burden of IHD among older adults has increased substantially ^3^, with a corresponding increase in diagnostic evaluations. In the United States alone, more than 1.5 million stress nuclear imaging studies are performed annually in the Medicare population ^4,5^. Similarly, IHD in women remains an important cause of morbidity and mortality. Recent estimates suggest that more than half of all women who develop heart disease have underlying IHD, and women often present with atypical symptoms that complicate diagnosis ^6,7^. Accordingly, a substantial proportion of non-invasive cardiac imaging tests are performed in women, in part to clarify diagnostic uncertainty ^8^.

Although many randomized controlled trials (RCT) have informed the use of non-invasive imaging in diagnosis and management of IHD, it is unclear how generalizable these trials are to women and older adults ^9–11^. Historically, women and older adults have been excluded or are underrepresented in RCTs of cardiovascular drugs and devices ^12–14^, but less is known about their representation in RCTs specifically evaluating non-invasive imaging modalities. This gap is critical because IHD may present differently in these populations, leading to different imaging test characteristics and differing responses to imaging-guided treatment ^15^. Additionally, older adults more frequently have multimorbidity and frailty, which may make them vulnerable to complications from testing itself while modifying potential benefits from subsequent interventions ^16^. Meanwhile, ischemic symptoms due to microvascular involvement are more common in women, which is not always well-detected on standard imaging tests and may respond differently to invasive or medical therapies for IHD ^17^.

In this systematic review, we sought to assess the proportional representation of women and older adults relative to the prevalence of IHD in these populations, in RCTs of non-invasive imaging and imaging-guided management of IHD.

## Methods

### Search overview

A systematic search was conducted across PubMed, ClinicalTrials.gov, and clinical guidelines of chest pain and chronic coronary syndrome to identify RCTs evaluating non-invasive imaging and imaging-guided management for acute and stable chest pain and IHD. On PubMed, filters were applied to include only RCTs published in English, involving human subjects aged 19 years and older, and published between January 1, 2002, and December 31, 2023. The search utilized the terms “chest pain” and “cardiac imaging” Studies were excluded based on the following criteria: non-randomized designs (non-randomized clinical trials, prospective cohorts, observational studies, validation studies, secondary analyses), studies of invasive procedures in one of the study arm without a corresponding non-invasive cardiac imaging comparator, not related to cardiac chest pain or cardiac imaging, rationale and design manuscripts, study protocols and studies terminated early due to poor enrollment.

The ClinicalTrials.gov search was conducted using the search terms “coronary artery disease” (for condition/disease) and “diagnostic testing” (for other terms), “chest pain” and “cardiac imaging” or “non-invasive imaging”, with filters for study status (completed or terminated) and study type (interventional, excluding observational studies), covering studies conducted between January 1, 2002, and December 31, 2023, and participants aged 18 years and older. Additionally, references from clinical guidelines and recent review articles were screened ^18,19^. (**Figure 1**)

**Figure 1:**
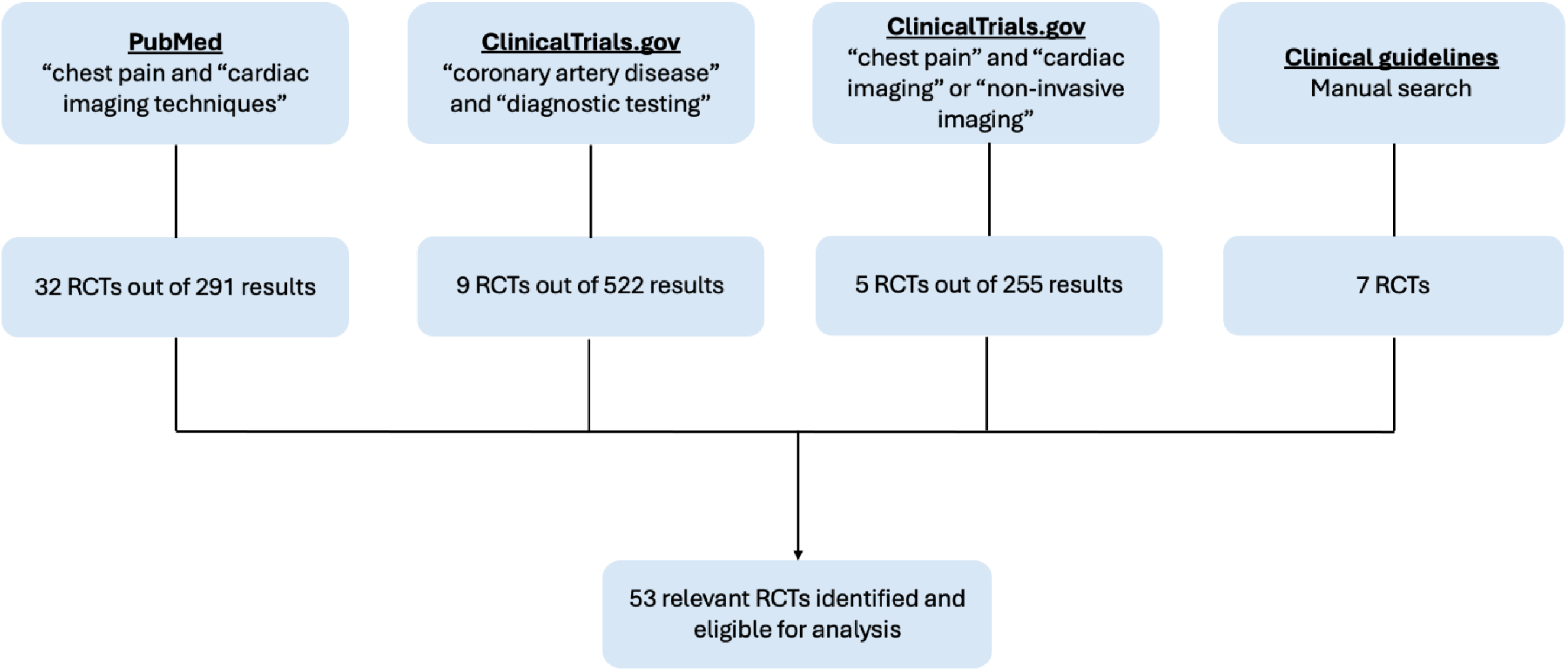
Flow-chart depicting study selection process.

### Data extraction and synthesis

Sex breakdown was available for all eligible RCTs. Age breakdown was available for 21 of the identified RCTs. When age subgroup breakdowns were not available in published reports, the first and/or corresponding author was contacted twice over three months to request data on age distribution.

### Primary outcome: Participation-to-Prevalence Ratio (PPR)

The primary outcome to quantitatively assess trial diversity was the Participation-to-Prevalence Ratio (PPR), which estimates the proportion of females and older adults in RCTs relative to their disease burden in the general population. The disease burden used for the denominators in the PPR calculations were derived from the Global Burden of Disease (GBD) study ^20^, which provides comprehensive data on mortality and disability trends worldwide, stratified by health condition, country, year, age, and sex. For single-country trials, the country-specific prevalence of IHD was used; for multinational trials, global prevalence estimates were used.

The PPR was calculated for each relevant subgroup (women, and age <65, 65–74, and ≥75 years) by dividing the proportion of trial participants in that subgroup by the proportion of that subgroup in the broader population with IHD ^21–23^. The interpretation of PPR was based on threshold in prior studies: PPR <0.8, 0.8-1.2, and >0.8 indicates underrepresentation, appropriate representation, and overrepresentation ^21,24^.

### Analysis

For each eligible trial, the PPR was calculated for women and across age groups of <65 years, 65–74 years, and ≥75 years as described above. Subgroup analysis was conducted for acute vs. stable chest pain population; and for trials performed in United States (US) vs. non-US to evaluate for any subgroup differences. An independent-samples t-test or Wilcoxon rank-sum test (for non-normal distributions) was performed to evaluate differences in mean PPR values across age groups (<65, 65–74, ≥75 years) and for female enrollment for the subgroup analyses.

## Results

We identified 53 eligible trials representing 55,893 participants. All 53 RCTs provided data on enrollment of women, while 21 RCTs representing 35,503 participants provided data by age subgroups (<65, 65-74, ≥75 years) **(Table 1)** ^25–78^. The lack of age subgroup data in the remaining trials was due to non-responsiveness from the corresponding or first author or unavailability of original trial data in older studies. Of these 53 trials, 25 enrolled patients with acute chest pain, while 28 enrolled patients with stable chest pain (**Table 2**). In terms of geographic distribution, 21 studies were conducted in the US (39.6%), 11 in the United Kingdom (20.8%), 4 in the Netherlands (7.6%), 2 in Germany and Denmark (3.8%), 1 each in Australia, Singapore, South Korea, China, and Canada (1.9%), and 8 in multiple countries (15.1%).

**Table 1:**
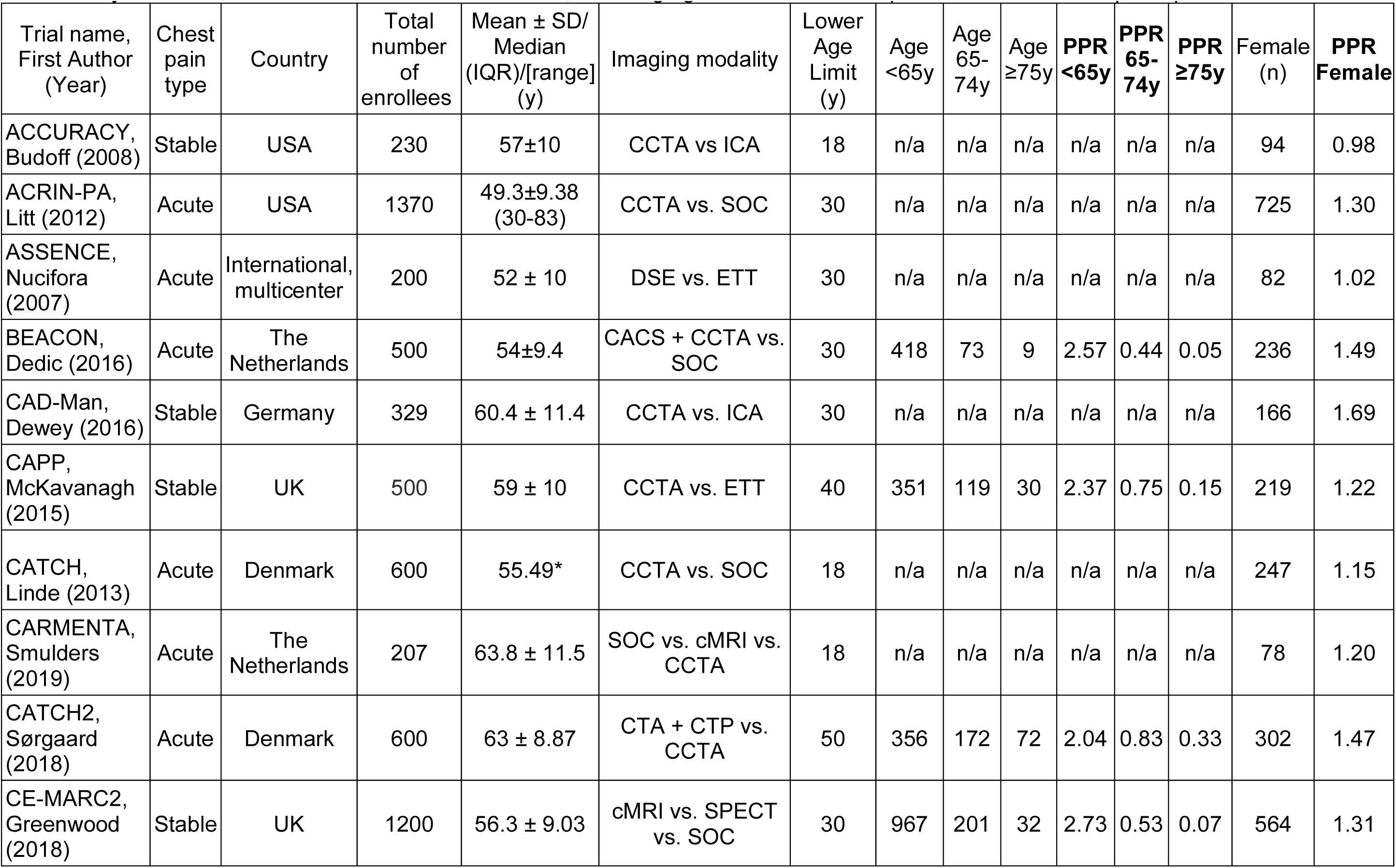
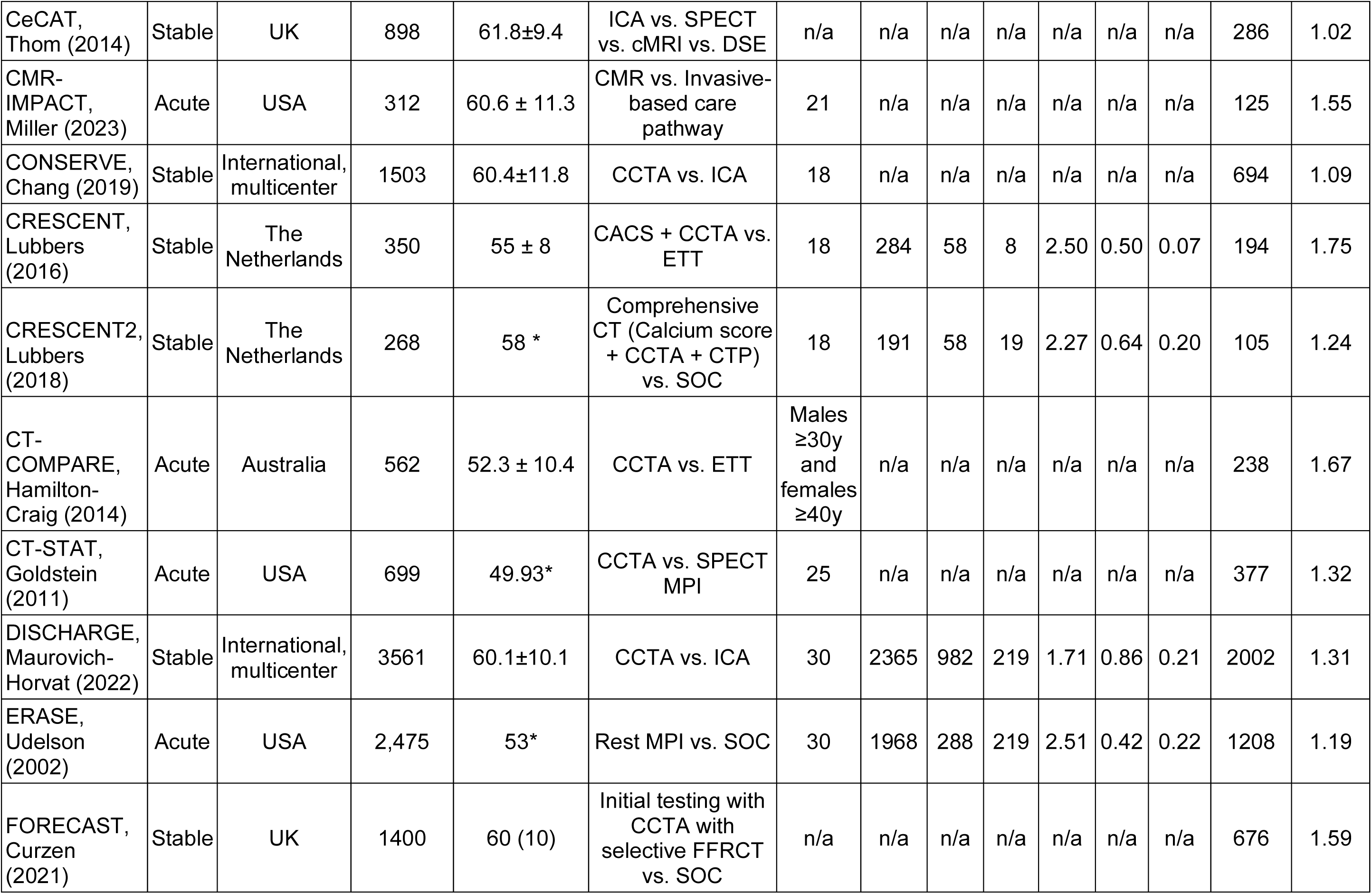
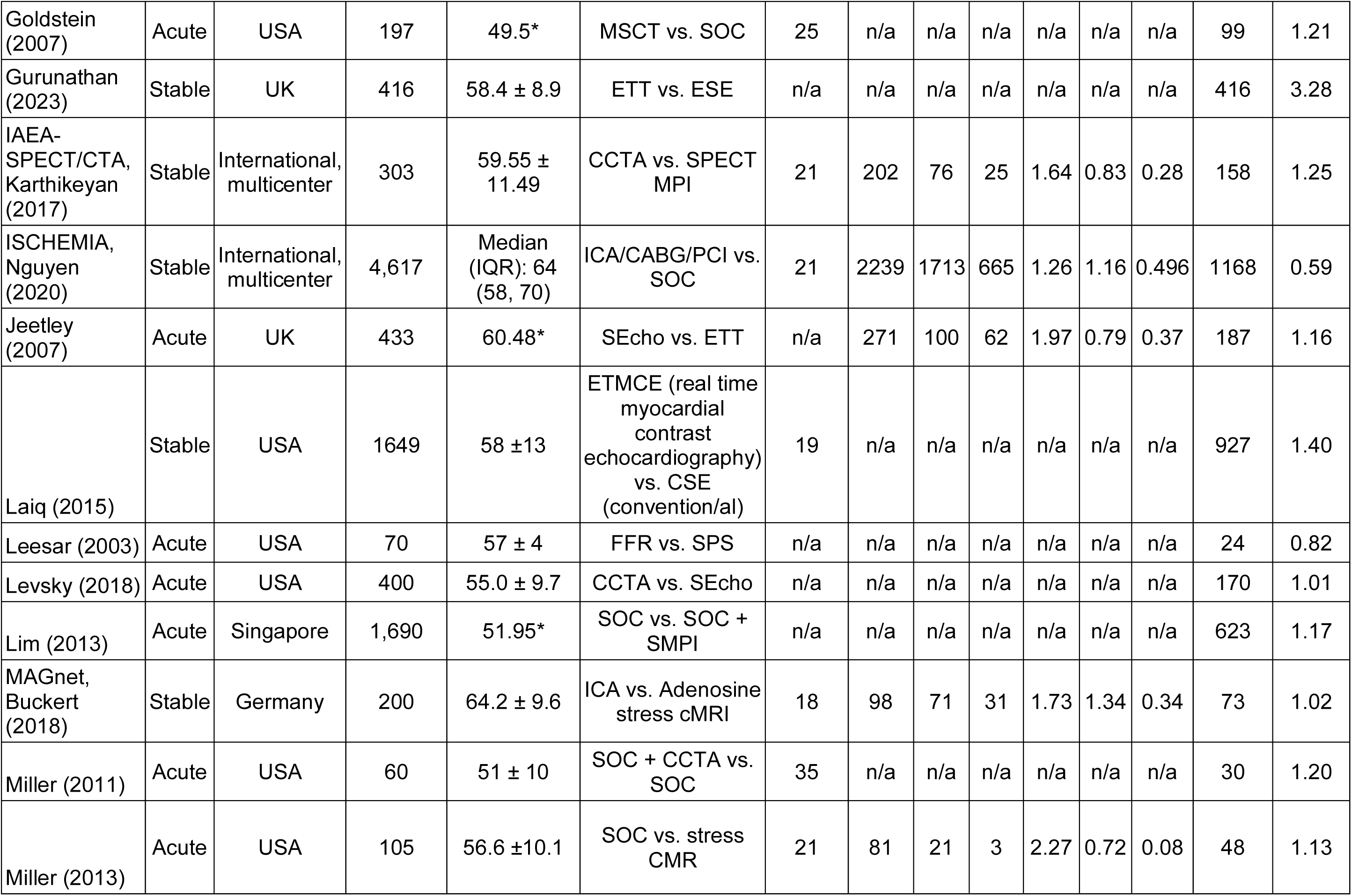
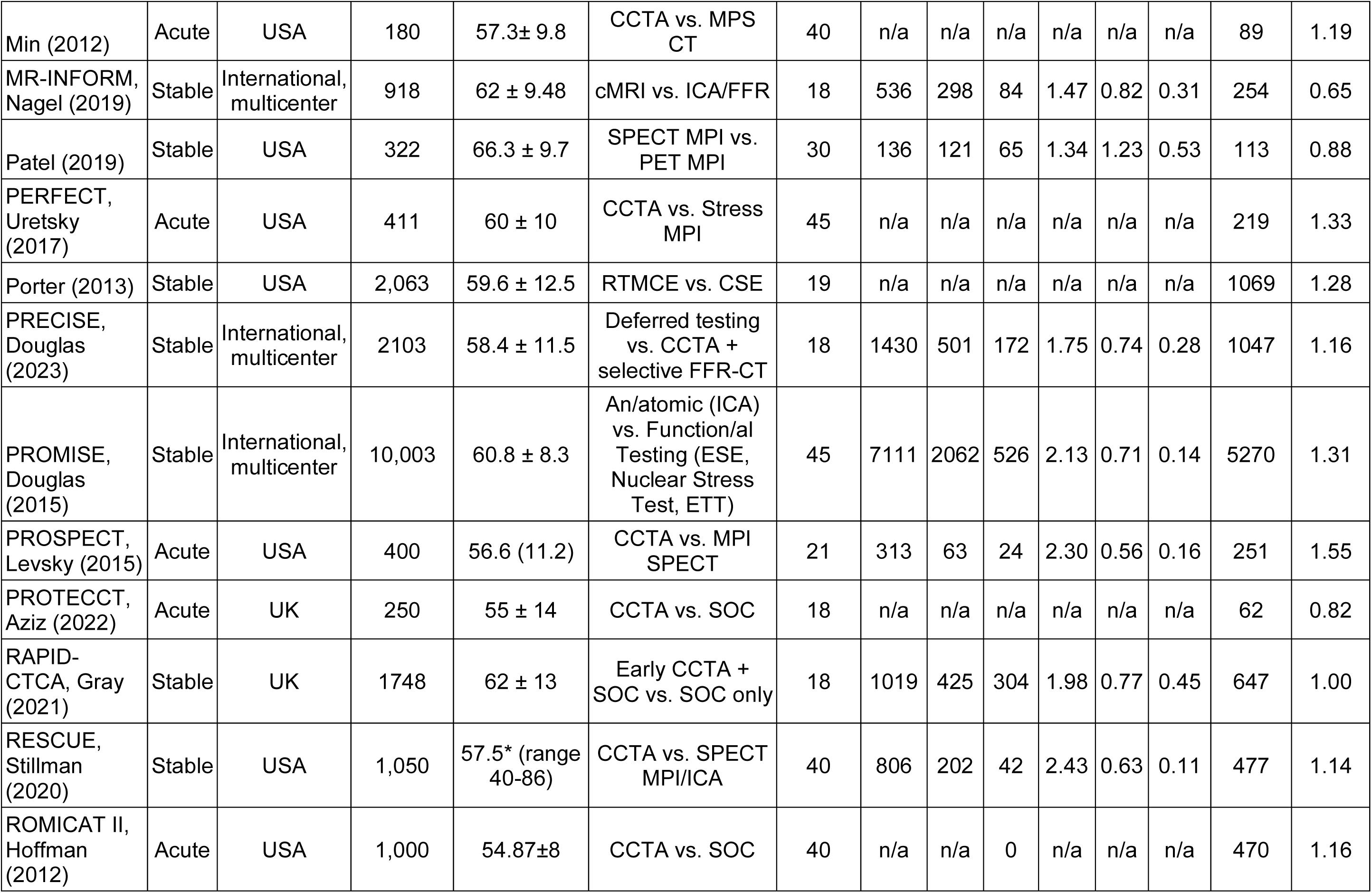
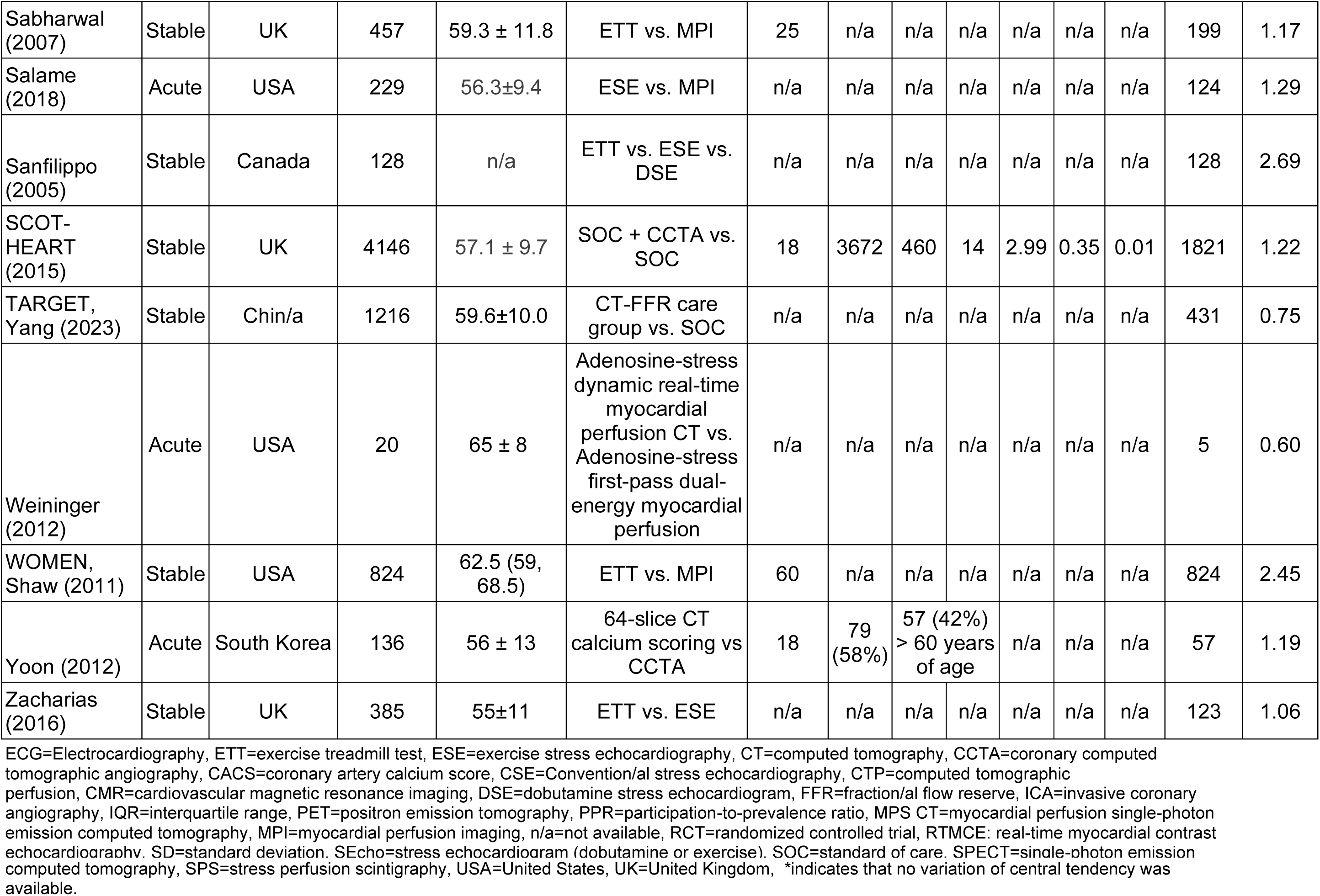
Key information from randomized controlled trials, including age distribution and representation of female participants.

**Table 2.** Overview of data from eligible randomized controlled trials.

| **Randomized controlled trials (RCTs)** | **n (%)** |
| --- | --- |
| Available age data | 52 (98) |
| Available age data by subgroups | 21 (40) |
| Available gender data | 53 (100) |
| Acute chest pain | 25 (35) |
| Stable chest pain | 23 (65) |
| Mean/median age <65 years | 51 (96) |
| Mean/median age 65-74 years | 1 (2) |
| Mean/median age ≥75 years | 0 |
| **Participation-to-prevalence ratio (PPR)** | **Median (interquartile range)** |
| Age <65 years | 2.13 [1.73, 2.43] |
| Age 65-74 years | 0.74 [0.56, 0.83] |
| Age 75 years | 0.21 [0.11, 0.33] |
| Female | 1.20 [1.06, 1.32] |
| **Trial by country** | **n (%)** |
| United States of America | 21 (39.6) |
| United Kingdom | 11 (20.7) |
| Netherlands | 4 (7.5) |
| Germany | 2 (3.8) |
| Denmark | 2 (3.8) |
| Australia | 1 (1.9) |
| Canada | 1 (1.9) |
| China | 1 (1.9) |
| Singapore | 1 (1.9) |
| South Korea | 1 (1.9) |
| International multicenter trials | 8 (15.1) |

The median age across all trials was 57.4 years [interquartile range, IQR: 55.0, 60.2] (**Table 2**). Overall, age was reported as a mean ± standard deviation (SD) in 41 trials while 9 trials reported overall age as median (range) or median (IQR). The mean/median age of all the trials ranged between 48 and 66 years (**Table 2**). No trials exclusively enrolled patients aged ≥75 years. Among the 21 studies with age breakdowns (<65 years, 65-74 years, and ≥75 years), 24,814 participants (69.9%) fell within the <65-year category, 8,064 (22.7%) in the 65-74-year category, and 2,625 (7.4%) in the ≥75-year category (**Central Illustration**).

The median PPR [IQR] by age across the 21 studies was 2.13 [1.73–2.43] for participants <65 years, 0.74 [0.56–0.83] for participants 65–74 years, and 0.21 [0.11–0.33] for participants ≥75 years (**Table 2**). Overall, older adults, particularly those ≥75 years, were underrepresented relative to the prevalence of IHD in these age groups, and younger adults <65 years were consistently overrepresented (**Figure 2**).

**Figure 2:**
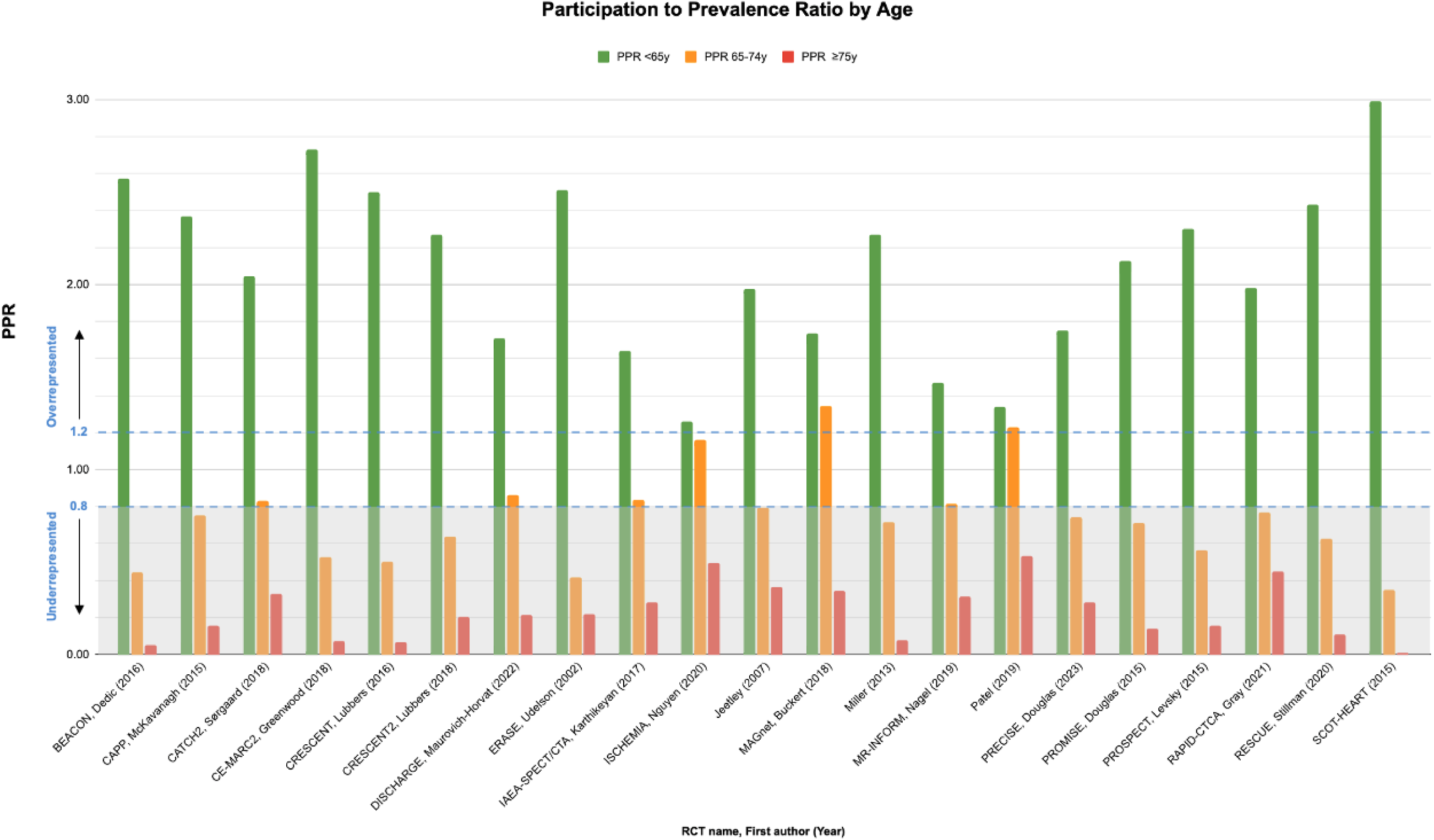
Participation-to-Prevalence Ratio (PPR) Across Three Age Groups from 21 out of 53 eligible trials with available age subgroup data. PPR <0.8, 0.8-1.2, >1.2 denotes under-representation, appropriate representation and over-representation respectively.

Enrollment data on women were available for all eligible trials. The median PPR [IQR] for women across all 53 studies was 1.2 [1.06–1.32]. Women were proportionally represented relative to the burden of IHD among women (**Figure 3**), with 49 trials (92.5%) demonstrating a PPR >0.8 for women. PPR results for both age and women representation were similar in US vs. non-US trials (**Figure 4**) and in acute versus stable chest pain trials (**Figure 5**).

**Figure 3:**
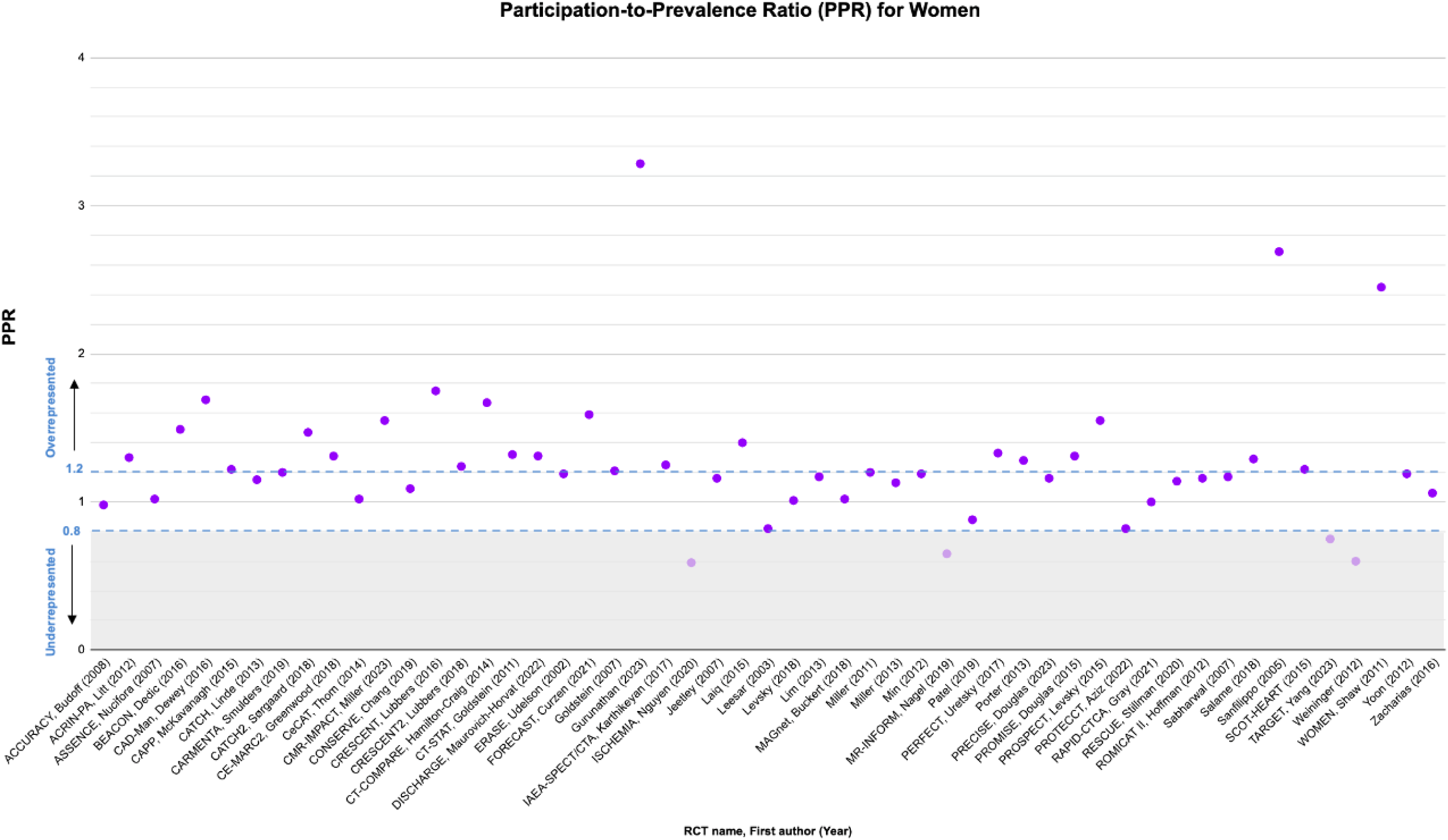
Participation-to-Prevalence Ratio (PPR) of Females Across 53 eligible Trials. PPR <0.8, 0.8-1.2, >1.2 denotes under-representation, appropriate representation and over-representation respectively.

**Figure 4.**
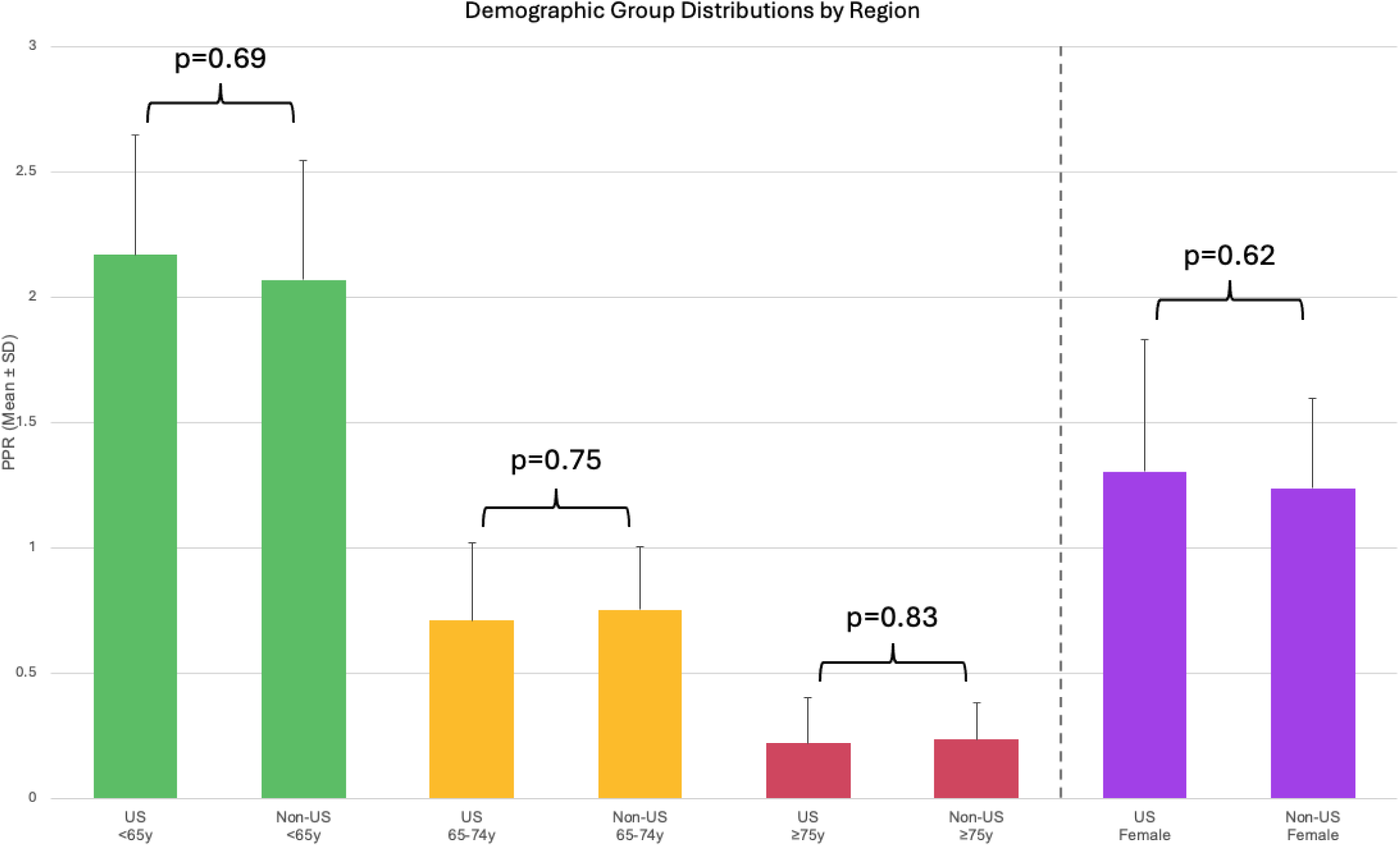
Demographic group distribution by trial region (US vs. non-US). PPR = Participation-to-prevalence ratio, US = United States. Age subgroup information available from 21/53 RCTs, sex information available from all 53 RCTs.

**Figure 5.**
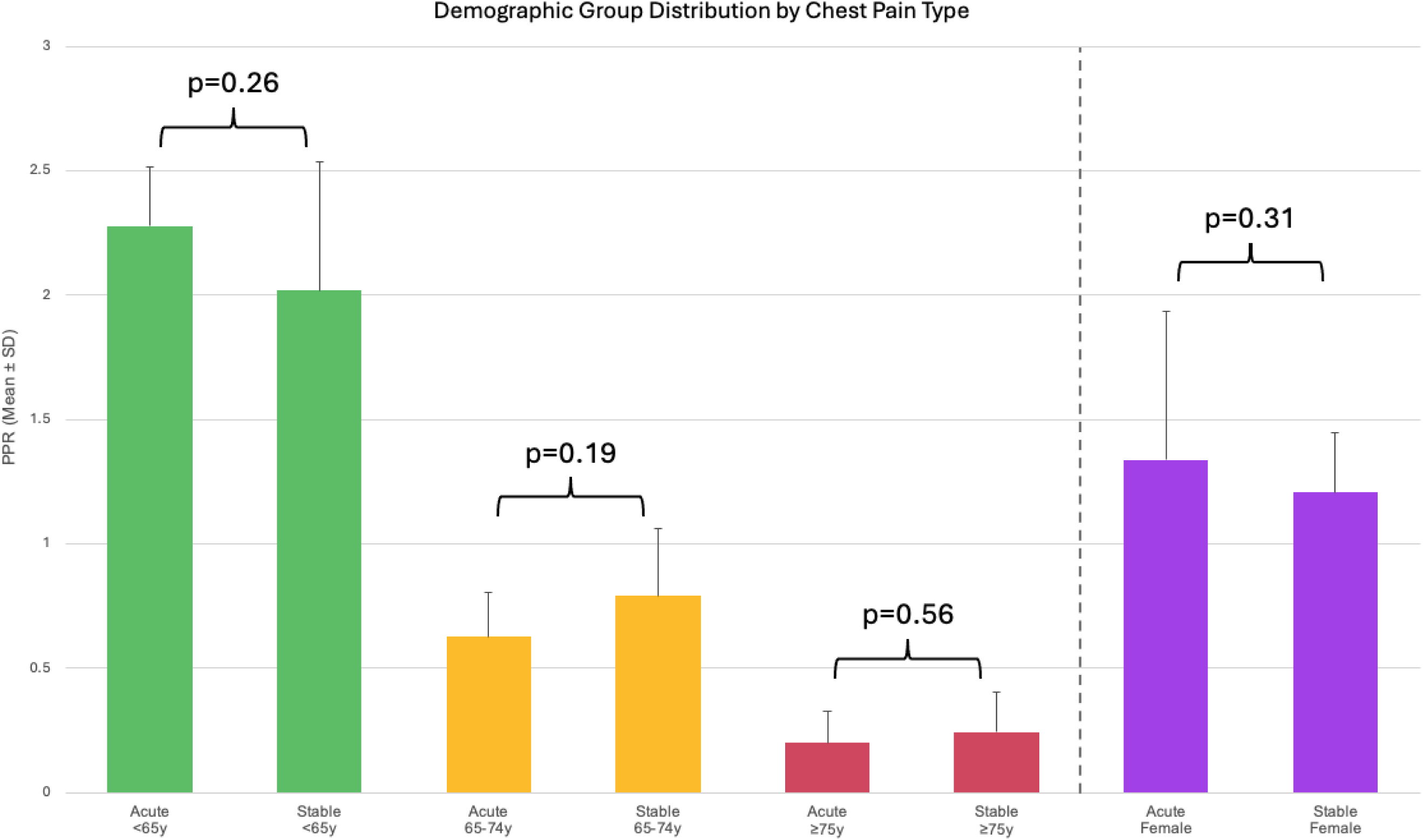
Demographic Group Distributions by Chest Pain Duration. PPR = Participation-to-prevalence ratio. Age subgroup information available from 21/53 RCTs, sex information available from all 53 RCTs.

## Discussion

In this systematic review, there was notable underrepresentation of adults aged 65–74 years and ≥75 years in RCTs evaluating non-invasive imaging-based diagnosis and management of IHD. By contrast, women were appropriately represented, with most trials meeting or exceeding the threshold for appropriate representation based on the PPR. These findings show that despite widespread reliance on non-invasive imaging for IHD management in older adults, RCTs generally do not sufficiently include older adults in numbers proportional to their disease burden. This study is one of the first to apply the PPR metric specifically to non-invasive imaging RCTs in patients with suspected or confirmed IHD, offering a more direct assessment of the generalizability of trials to these underrepresented populations.

There are several potential explanations for the underrepresentation of patients >75 years in clinical trials of imaging for IHD. Older adults frequently experience multimorbidity and frailty ^79^, which may disqualify them from RCT participation or discourage recruitment due to concerns about comorbid conditions, polypharmacy, and higher procedural risk ^80^. Additionally, many trial protocols have upper age limits or exclude individuals with complex comorbidities ^31,34,81^. Even when older adults are enrolled, investigators may face challenges regarding informed consent and retention ^82^. However, the absence of robust data in the elderly impedes understanding of how various imaging strategies performed in the setting of geriatric conditions such as frailty, cognitive impairment, or multimorbidity commonly present in older adults.

This study also demonstrates that women achieved near-equitable representation in most imaging RCTs relative to the underlying prevalence of IHD. This finding contrasts with reports and systematic reviews from other areas of cardiovascular research, where women have often been underrepresented ^8,83^. This may reflect growing awareness of sex-based disparities, along with regulatory efforts and guidelines encouraging or requiring sex-specific reporting and even the including sex analysis in clinical trial planning ^84^. Nonetheless, although women are proportionally included overall, additional research is warranted to examine whether imaging modalities are equally effective in women - particularly those with microvascular disease or seemingly non-cardiac presentations.

Underrepresentation of older adults in RCTs limits the generalizability of trial findings to real-world practice, where comorbidities and frailty can alter both diagnostic performance and clinical outcomes. Future trial designs should emphasize the inclusion of older adults, potentially by expanding eligibility criteria or designing trials specifically to evaluate effectiveness in older adults, adopting adaptive designs that allow for subgroup analyses, and addressing logistical barriers such as transportation and caregiver support. In addition, further evaluation of potential sex-specific differences in imaging test accuracy and subsequent management is needed, even as overall representation of women may improve.

### Strengths and Limitations

A key strength of this review is its focus on RCTs specifically evaluating non-invasive imaging strategies, which are central to contemporary IHD management. We employed a comprehensive search strategy, including clinical trial registries and guideline references, and systematically evaluated the age and sex distribution of trial participants using a validated metric. Limitations include potential publication bias and incomplete reporting of age subgroup data. Although we attempted to contact authors for missing age data, not all authors responded. Furthermore, the PPR depends on accurate population prevalence estimates, which may not perfectly reflect the patient mix in all regions or time periods.

## Conclusion

RCTs of non-invasive imaging in IHD appear to adequately represent women but consistently underrepresent older adults, especially those ≥75 years of age. Future trials must prioritize enrolling older adults to ensure that clinical recommendations drawn from these studies are relevant to the patients most likely to undergo imaging and benefit from evidence-based care.

## Data Availability

The study involves the analysis of publicly accessible, de-identified data and therefore does not require IRB review as it does not constitute human subjects research under federal regulations.

